# Host genetic liability for severe COVID-19 overlaps with alcohol drinking behavior and diabetic outcomes in over 1 million participants

**DOI:** 10.1101/2020.11.08.20227884

**Authors:** Frank R Wendt, Antonella De Lillo, Gita A Pathak, Flavio De Angelis, COVID-19 Host Genetics Initiative, Renato Polimanti

## Abstract

To distinguish correlation from causation, we performed *in-silico* analyses of three COVID-19 outcomes (N>1,000,000). We show genetic correlation and putative causality with depressive symptoms, metformin use, and alcohol use. COVID-19 risk loci associated with several hematologic biomarkers. Comprehensive findings inform genetic contributions to COVID-19 epidemiology, molecular mechanisms, and risk factors.

## Main Text

Host genetic liability and epidemiologic risk for severe COVID-19 (coronavirus disease 2019) following SARS-Cov-2 (severe acute respiratory syndrome coronavirus 2) infection is of immediate clinical interest [1]. Genome-wide association studies (GWAS) of COVID-19 outcomes identified several risk loci and provided evidence of the pleiotropic effects (i.e., a single locus confers risk to several similar or disparately related traits) shared with other human diseases and traits [2]. To understand better the association of biological measurements, lifestyle indicators, biomarkers, and health and medical records with COVID-19 susceptibility, we performed analyses to distinguish genetic correlation (e.g., shared genetic liability) from genetically-informative putative causal effects. These results provide insights into the mechanisms underlying several associations linking COVID-19 susceptibility to human diseases and traits.

## Methods

Genome-wide association statistics were accessed from the COVID-19 Host Genetics Initiative (HGI; https://www.covid19hg.org/; September 30, 2020 release date) [3] for seven COVID-19 phenotypes. Using Linkage Disequlibrium Score Regression (LDSC [4]), we calculated common variant heritability (h^2^). Three outcomes presented with SNP-based h^2^ z-scores > 0: *very severe respiratory confirmed COVID-19 versus population* (COVID_19_A2; N_case_=2,972, N_control_=284,472), *hospitalized COVID-19 versus COVID-19 population* (COVID_19_B2; N_case_=6,942, N_control_=1,012,809), and *COVID-19 versus population* (COVID_19_C2; N_case_=17,607, N_control_=1,345,334). The h^2^ estimates of four other COVID-19 outcomes curated by HGI were not significantly different from 0 (Table S1).

For two COVID-19 phenotypes with h^2^ z-scores > 4, *severe respiratory COVID-19* and *hospitalized COVID-19*, we then estimated their genetic correlation (rg) with 4,083 phenotypes from the UK Biobank (UKB, see http://www.nealelab.is/uk-biobank). LDSC analyses were based on linkage disequilibrium information from the 1000 Genomes Project (1kGP) European reference population. For continuous traits, we restricted our analyses to genome-wide association statistics generated from inverse-rank normalized phenotypes.

To distinguish between genetic correlation and causative effects, we applied the Latent Causal Variable (LCV) approach [5]. Under the assumption of a single effect-size distribution in per-trait GWAS, LCV tests for the presence of a single latent trait connecting COVID-19 outcomes to UKB phenotypes. LCV was performed in R using the 1kGP European reference LD panel and genome-wide association statistics for SNPs with minor allele frequencies >5%. Variants in the major histocompatibility complex region of the genome were excluded because of its complex LD structure. LCV gĉp estimates were only interpreted for trait pairs where both traits exhibit LCV-calculated h^2^ z-scores ≥ 7. The gĉp estimate ranges from 0-1 with values near zero indicating partial causality and values approaching 1 indicating full causality. LCV developers recommend that gĉp>0.7 is evidence of a fully causal relationship between trait pairs [5].

Phenome-wide association studies (PheWAS) were performed for 14 loci associated with one of the three heritable COVID-19 outcomes (Table S2) using the Pan-ancestry UK Biobank resource (available at https://pan.ukbb.broadinstitute.org/downloads). We analyzed genome-wide association statistics generated from the analysis of 7,218 phenotypes in six ancestries: European (N=420,531), Centra/South Asian (N=8,876), African (N=6.636), East Asian (N=2,709), Middle Eastern (N=1,599), and Admixed American (N=980). Pan-UKB traits were analyzed if they had 100 cases in European ancestry or 50 cases in all other ancestries. Association statistics were covaried with sex, age, age^2^, sex×age, sex×age^2^, and the first ten within-ancestry principle components. A detailed description of the methods used to generate these data is available at https://pan.ukbb.broadinstitute.org/. Multiple testing correction was performed for the number of phenotypes and ancestry groups using the p.adjust(method= ‘fdr’) function of R.

## Results

Assuming a population prevalence of 1%, the h^2^ of *severe respiratory COVID-19* (A2 h^2^=0.129, se=0.024, p=5.95×10^−8^) and *hospitalized COVID-19* (B2 h^2^=0.0411, se=0.010, p=2.74×10^−5^) were significantly different from zero. The h^2^ for *COVID-19 versus population* was significantly different from zero (C2 h^2^=0.0089, se=0.003, p=0.007) but not accurate enough for genetic correlation. *Severe respiratory COVID-19* and *hospitalized COVID-19* were genetically correlated with 127 and 174 phenotypes, respectively, after multiple testing correction (Figure 1A). The most significant genetic correlate of each phenotype was waist circumference (*severe respiratory COVID-19* r_g_=0.272, se=0.046, p=2.18×10^−9^ and *hospitalized COVID-19* r_g_=0.342, se=0.061, p=1.66×10^−8^). Five phenotypes were significantly genetically correlated with *severe respiratory COVID-19* after multiple testing correction and not correlated (unadjusted p-value >0.05) with *hospitalized COVID-19* (Table S3) The strongest of these was the genetic correlation between UKB Field ID 20110_100 “lack of maternal history of heart disease, stroke, high blood pressure, chronic bronchitis/emphysema, Alzheimer’s disease/dementia, diabetes” and *severe respiratory COVID-19* (r_g_=-0.231, se=0.078, p=0.003; versus *hospitalized COVID-19* r_g_=-0.119, se=0.099, p=0.234).

**Figure 1.**
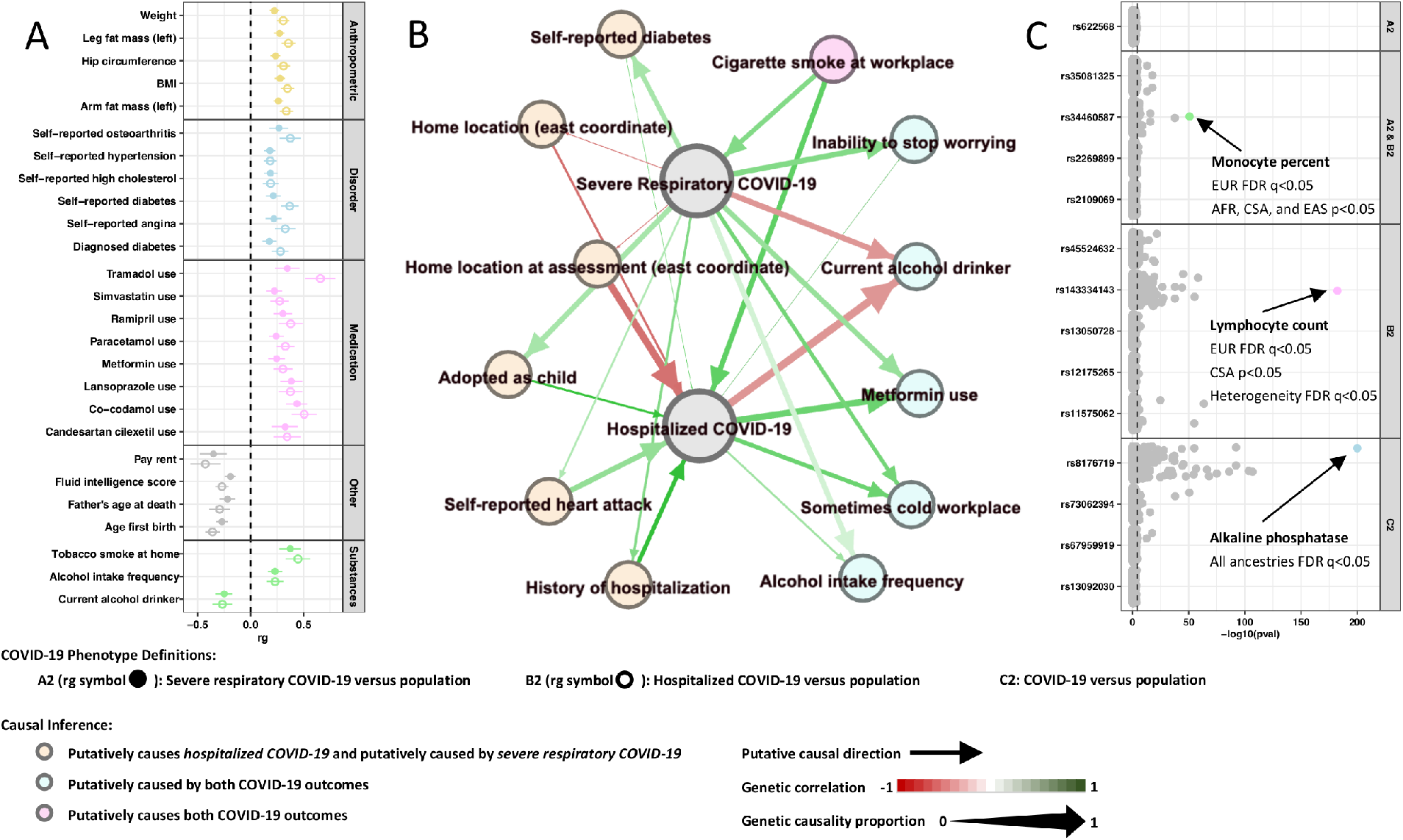
Pleiotropy of genetic risk for COVID-19 susceptibility. (A) all nominally significant genetic correlations; (B) latent causal variable network for phenotypes meeting FDR significance in A2 and B2; (C) PheWAS results.

With 188 traits genetically correlated with either COVID-19 outcome after multiple testing correction (Table S3), we tested for causality among UKB, *severe respiratory COVID-19, hospitalized COVID-19*. After multiple testing correction we detected 24 and 42 latent causal genetic relationships with *severe respiratory COVID-19* and *hospitalized COVID-19*, respectively (Table S4). *Severe respiratory COVID-19* showed a partial casual effect for manifestations of mania or irritability (gĉp=0.685, se=0.202, p=0.5.14×10^−4^) and candesartan cilexetil use (gĉp=0.641, se=0.374, p=2.30×10^−7^). *Hospitalized COVID-19* was fully genetically causal for diabetes (gĉp=0.831, se=0.289, p=9.42×10^−7^) and alcohol drinking status (gĉp=0.848, se=0.089, p=4.13×10^−13^). Twelve phenotypes had significant causal estimates with both *severe respiratory COVID-19* and *hospitalized COVID-19* (Figure 1B). These included smoking and drinking behaviors, diabetes, and heart attack. After multiple testing correction there were no significant differences between genetic causality proportions estimated for *severe respiratory COVID-19* and *hospitalized COVID-19*.

Phenome-wide assessment of 14 COVID-19 liability loci (across three severity outcomes: *severe respiratory, hospitalized COVID-19*, and *all COVID-19*) identified 439 significant (FDR q<0.05, Figure 1C) out of 7,221 phenotypes across six ancestries (Table S5). After multiple testing correction, alkaline phosphatase was significantly negatively associated with rs8176719 (*ABO* locus) in all six ancestries. The *ABO* locus exhibited significant effect size heterogeneity (Figure S1) with respect to low-density lipoprotein cholesterol concentration (cross-ancestry meta-analysis β=0.057, p=3.01×10^−5^) and hemoglobin concentration (cross-ancestry meta-analysis β=0.040, p=1.67×10^−5^). Outside the *ABO* locus, we detected significant effect size heterogeneity at rs143334143 (*CCHCR1* locus, lymphocyte count meta-analysis β=0.144, p=5.16×10^−179^, p_het_=2.95×10^−5^) and rs45524632 (*KEAP1* locus, heel bone mineral density T-score meta-analysis β=-0.042, p=2.59×10^−5^, p_het_<2.95×10^−5^).

## Discussion

To date, millions of people worldwide have been infected with SARS-CoV-2, which is the causative agent responsible for global lockdowns and heavily restricted interpersonal contact after widespread COVID-19 outbreak. In light of the 2020 COVID-19 pandemic, host genetic susceptibility to severe responses to SARS-CoV-2 infection is critical. Until recently, genetic epidemiologic approaches to understand host susceptibility have been limited due to relatively small sample sizes and reduced statistical power. In this investigation we use genome-wide data to uncover overlap and putative causal relationships between genetic liability to COVID-19 severity, preclinical risk factors (e.g., alcohol consumption) [6], and long-term consequences of infection (e.g., diabetes) [7]. These data reflect potential measures to refine, and/or improve accuracy and generalizability of, COVID-19 severity outcomes with epidemiological and self-report information [1].

Our most notable findings reflect (1) causal consequences of cigarette smoke exposure on COVID-19, (2) causal consequences of COVID-19 on diabetes, and (3) ABO blood type effects on COVID-19 severity across ancestry [8]. Alcohol and diabetes relationships with COVID-19 severity demonstrate that epidemiologic relationships between them are due to putative causal effects [9]. Persons with diabetes have been identified as some of the most high risk individuals and there are several instances of spontaneous diabetes onset following COVID-19 recovery [10]. To our knowledge, this is the first indication that genetic liability to COVID-19 severity also contributes to diabetes at levels suggesting a “fully causal” relationship. With single-SNP measures we recapitulate the relationship between COVID-19 severity and diabetes outcomes by detecting consistent negative association between rs8176719 (*ABO* locus) and alkaline phosphatase, an enzyme with evidence of protective effects against diabetes when present in sufficient concentrations [11].

Given the extremely large number of people infected with SARS-CoV-2 globally, these risk factors and potential chronic outcomes have critical public health consequences on the long-term economic burden of the COVID-19 pandemic [12].

## Data Availability

All data are available for public download at PanUKB (https://pan.ukbb.broadinstitute.org/), HGI (https://www.covid19hg.org/), UKB (http://www.nealelab.is/uk-biobank).

## Acknowledgements

The authors thanks the participants and the investigators involved in the COVID-19 Host Genetics Initiative.

## Supplementary Information

**Table S1**. Heritability (h^2^) estimates for seven COVID-19 outcomes in the COVID-19 Host Genetics Initiative (https://www.covid19hg.org/; September 30, 2020 release date).

**Table S2**. Genome-wide significant SNPs associated with COVID-19 outcomes.

**Table S3**. Significant genetic correlations (rg; FDR q<0.05) between COVID-19 outcomes and phenotypes studied in the UK Biobank European ancestry cohort. A2: very severe respiratory COVID-19 versus population; B2: hospitalized COVID-19 versus population.

**Table S4**. Latent causal variable results (FDR q<0.05) to detect putative causal relationships between COVID-19 outcomes and phenotypes ascertained in the UK Biobank European ancestry cohort. A2: very severe respiratory COVID-19 versus population; B2: hospitalized COVID-19 versus population.

**Table S5**. Significant genome-wide association results (FDR q<0.05 in at least one ancestry) for COVID-19 risk loci detected across three phenotype definitions and six ancestries from the Pan-UKBiobank.

**Figure S1.**
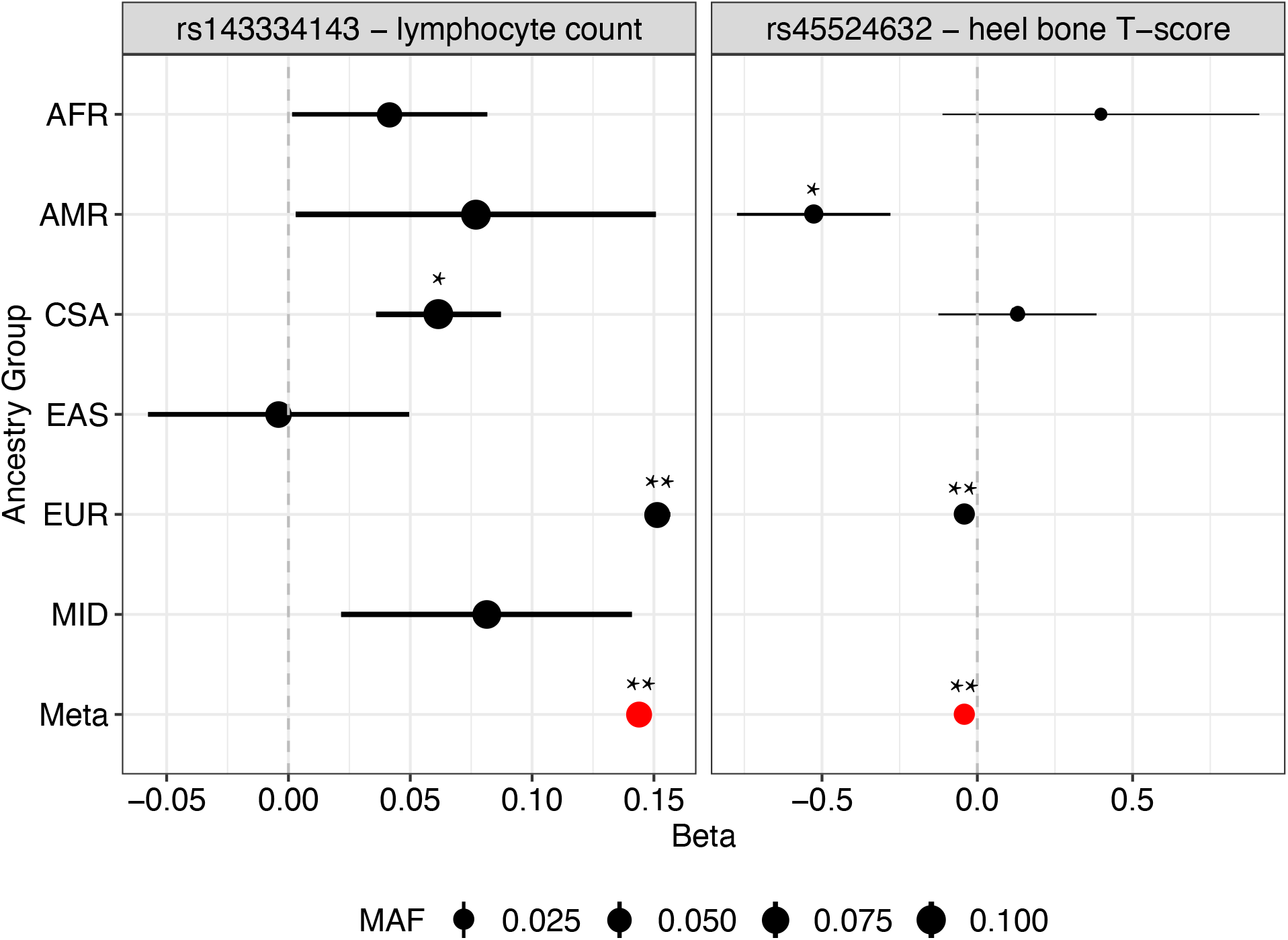
Heterogeneous effects of rs143334143 on lymphocyte count and rs45524632 on heel bone T-score across six world populations from the Pan-UK Biobank. Double asterisks indicate significance after multiple testing correction (FDR 5%) while single asterisks indicate nominal significance (p<0.05).

